# Data Management Plan for Healthcare: Following FAIR Principles and Addressing Cybersecurity Aspects. A Systematic Review using InstructGPT

**DOI:** 10.1101/2023.04.21.23288932

**Authors:** Alexandru Stanciu

## Abstract

This study focuses on data-related aspects and emphasizes the importance of producing a data management plan (DMP) to address the challenges specific to the collection, processing, storage, security, documentation, sharing, and distribution of data in research projects in the healthcare sector. It provides an overview of the DMP and offers guidelines for creating an effective plan that incorporates the FAIR principles. Additionally, the study outlines the main aspects of data management in the healthcare domain and analyzes several security issues, such as cryptography, biometrics, and digital watermarking, that should be considered for healthcare data. A systematic review of the literature is performed to explore the critical aspects of data management and identify emerging trends, challenges, and innovative solutions that can be incorporated into DMPs. Part of the analysis of this survey was performed with the InstructGPT language model.

## 1 Introduction

One of the most critical issues that need to be addressed during the lifetime of a research project is related to data management. For example, aspects related to the storage and distribution of data must be treated systematically, and documenting how the data was generated, processed, and shared should be carefully considered, as one of the main objectives of the research activity is to disseminate the results as widely as possible.

Moreover, an essential aspect of data management is related to its security. Researchers need to be confident that the data they are working with is securely stored and that confidential information is protected and cannot be altered, destroyed, or otherwise compromised. Last but not least, the specific requirements of funding organizations should be taken into account, as well as the legal provisions related to data privacy.

In typical research projects, it is necessary to address these issues by producing and using a data management plan (DMP) that specifies how these data-related activities should be carried out (i.e., to describe how data will be generated (including tools and methods used), how it will be stored, presenting both data sharing and distribution (addressing licensing arrangements), and ensuring its security, confidentiality, and integrity.

Furthermore, the interdisciplinary nature of healthcare data management requires a good understanding of the different components involved in building applications and services that handle the generation, processing, analysis, and storage procedures, the best practices for their implementation and operation, as well as the challenges that could be faced. Similarly, the rapid advancement of technology and increasing complexity of healthcare data implies a continuous evaluation and adaptation of data management strategies.

This study provides a brief description of the DMP, including some guidelines for designing an efficient plan, considering the FAIR principles related to research data. In order to provide a broader picture of the scope of a DMP, we present several aspects related to data management in the healthcare domain, emphasizing the issues related to data security. In addition, a systematic review with the primary aim to explore the critical aspects of data management in healthcare domain is performed to identify emerging trends, challenges, and innovative solutions that can be incorporated into DMPs.

For this task, we have outlined the following objectives, specifically: (a) to understand the role of FAIR principles in developing an effective DMPs for the healthcare domain; (b) to analyze the security and privacy protection measures that should be considered for healthcare data; (c) to synthesize the current state of knowledge on DMPs for healthcare domain and provide recommendation for further research.

Consequently, the research questions to guide the investigation are the following: (a) how do the FAIR principles contribute to the design of effective DMPs in the healthcare domain? (b) how are address security concerns, such as data privacy, and access control in DMPs? (c) what are the emerging trends, challenges, and innovations in healthcare data management?

The paper is structured as follows. Section 2 analyzes the data management plan in the healthcare sector, highlighting the role of the FAIR principles and reviewing several security aspects. Section 3 describes the materials and methods used for the systematic literature review, and Section 4 presents the results obtained. The final section provides the conclusions of the study.

## 2 Data Management Plan in Healthcare Domain

Data management plans are written documents that accompany research proposals and provide information about the data used and produced during research activities. DMPs specify where the data will be stored, which licenses and constraints apply, and who should receive credit for the data. They serve as useful tools to help researchers manage their data, maintain its quality, and make it accessible and reusable even after the research project has concluded.

Typically, DMPs are required by funding organizations and institutions worldwide, and researchers create them using checklists and online questionnaires. DMPs offer several potential benefits for different stakeholders involved in the research process. These benefits extend from funders and legal experts to researchers and publishers, repository operators, and institutional administrators.

Funders, for instance, benefit from having structured information about who is producing the data and where the data will be deposited. This information can be provided in the DMP, enabling funders to monitor compliance through automated processes rather than relying on manual methods. By having easy access to this information, funders can ensure that researchers are adhering to best practices for data management and sharing. Legal experts can benefit from relevant DMP content being reused in patent applications. By having access to detailed information about the data and research methods from the outset of the project, legal experts can identify and address any legal issues that may arise later on. Researchers benefit from DMPs by enabling connections with experts throughout the research project for data management advice and support. DMPs can also serve as an important source of information on experiment design and implementation, providing a comprehensive record of the research process. Publishers can use DMPs to generate automatic data availability statements and properly link and cite articles, datasets, and other outputs. By having access to this information, publishers can ensure that data is properly credited and that articles and datasets are linked in a way that enables maximum visibility and impact. Repository operators benefit from DMPs by receiving information about costs, licenses, metadata requirements, and other important details related to data management and operations. This enables better planning for capacity and facilitates data preservation. Finally, institutional administrators can get a holistic view of the data used, processed, and created within the institution. This helps in better planning of resources needed to support data management infrastructure. By having a comprehensive understanding of research activity across the institution, administrators can identify areas of strength and weakness and allocate resources accordingly.

Efficient management of data requires accurate information across a variety of areas, including technical details, formats, infrastructure, and legal and ethical considerations surrounding data collection and reuse. Developing a DMP should be a collaborative effort involving stakeholders with expertise in their domains and adjacent areas of the data management ecosystem. By doing so, the right information can be provided and acted upon by others.

Certain information needed for a DMP may already exist electronically, and it would be beneficial to retrieve this information from appropriate sources after checking for consistency and quality assurance. To accomplish this, it is necessary to integrate systems and allow stakeholders to expose services that automate tasks, such as gathering administrative data, like affiliation, grant number, and contact information, from institutional databases such as Current Research Information Systems (CRIS) or Research Information Management (RIM) systems, to pre-fill the DMP (Miksa et al., 2019).

To create an effective plan, it is important to provide adequate information about the data that will be collected. The most important aspects include:

- Types: Different types of data could be collected, such as text, spreadsheets, software, images, audio files, and patient records.
- Sources: Data may come from human observation, laboratory and field instruments, experiments, simulations, and other studies. It’s important to clarify whether the data is proprietary, subject to restrictions, or related to human subjects.
- Volume: The total amount of data and number of files to be collected can impact data management activities.
- Data and file formats: It’s crucial to choose formats that are non-proprietary, based on open standards, and widely used in the scientific community. Comma Separated Values (CSV) is preferred over Excel (.xls, xlsx). Uncompressed, unencrypted data stored using standard character encodings such as UTF-16 are more likely to remain accessible in the long term.

In some cases, it may not be possible to determine the exact types, sources, volume, and formats of data in advance, and the plan should be updated iteratively. Therefore, it is recommended to treat the DMP as a dynamic document that should be reviewed and updated frequently, ideally on a quarterly basis. Assigning a team member to revise the plan and keeping track of changes in a revision history can help to ensure that the plan stays up-to-date and reflects any new protocols or policies.

Accordingly, the metadata would need to include additional information such as the time of creation, the purpose of the data, the person responsible for it, and its previous usage (including who used it, why, how, and when). Having this metadata would enable data analysts to reproduce previous experiments and assist in future scientific studies.

A thorough data management plan (DMP) clearly outlines the duties and obligations of all individuals and organizations involved in the project (Michener, 2015). These responsibilities may include collecting, entering, quality checking, creating and managing metadata, backing up, preparing and submitting data to an archive, and administering systems.

To illustrate these aspects, we conducted a survey on a set of DMPs that were created within the context of research projects in the healthcare sector. In this regard, we identified the elements relevant to the implementation of FAIR principles, as well as the approach taken towards addressing issues related to data security and privacy. The results are presented in Table 1.

**Table 1:**
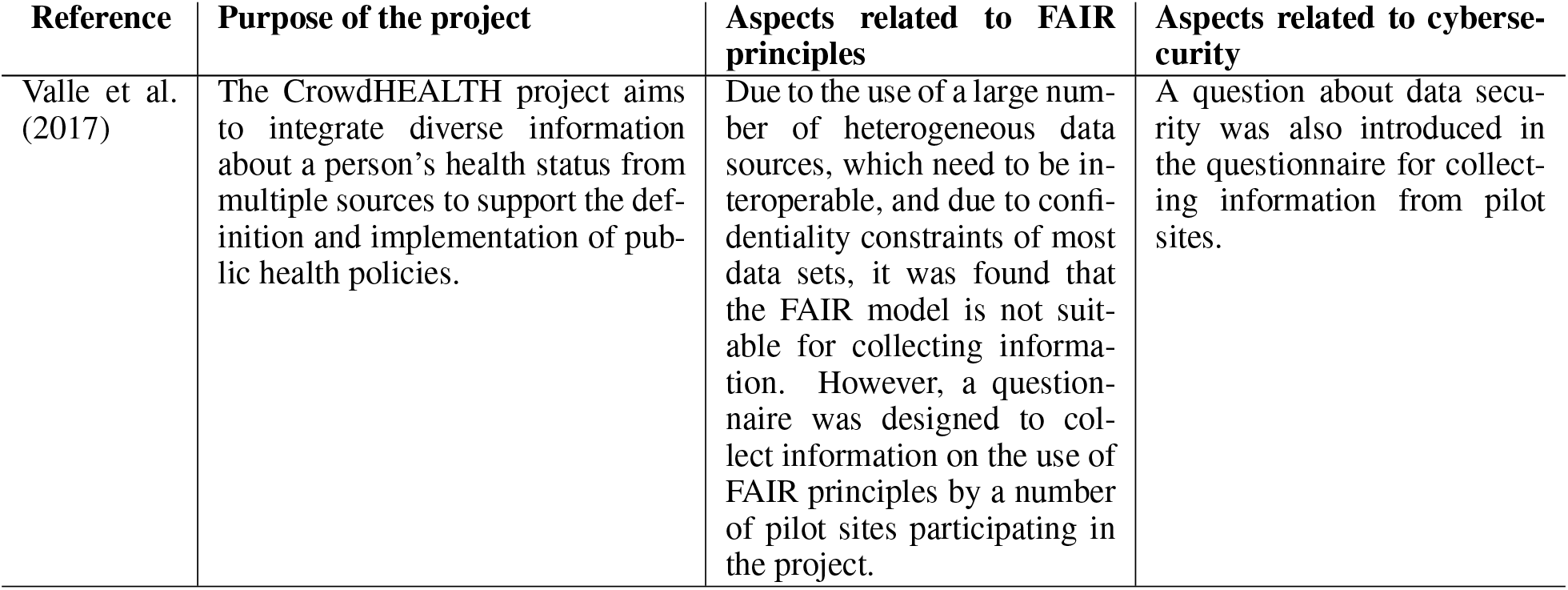
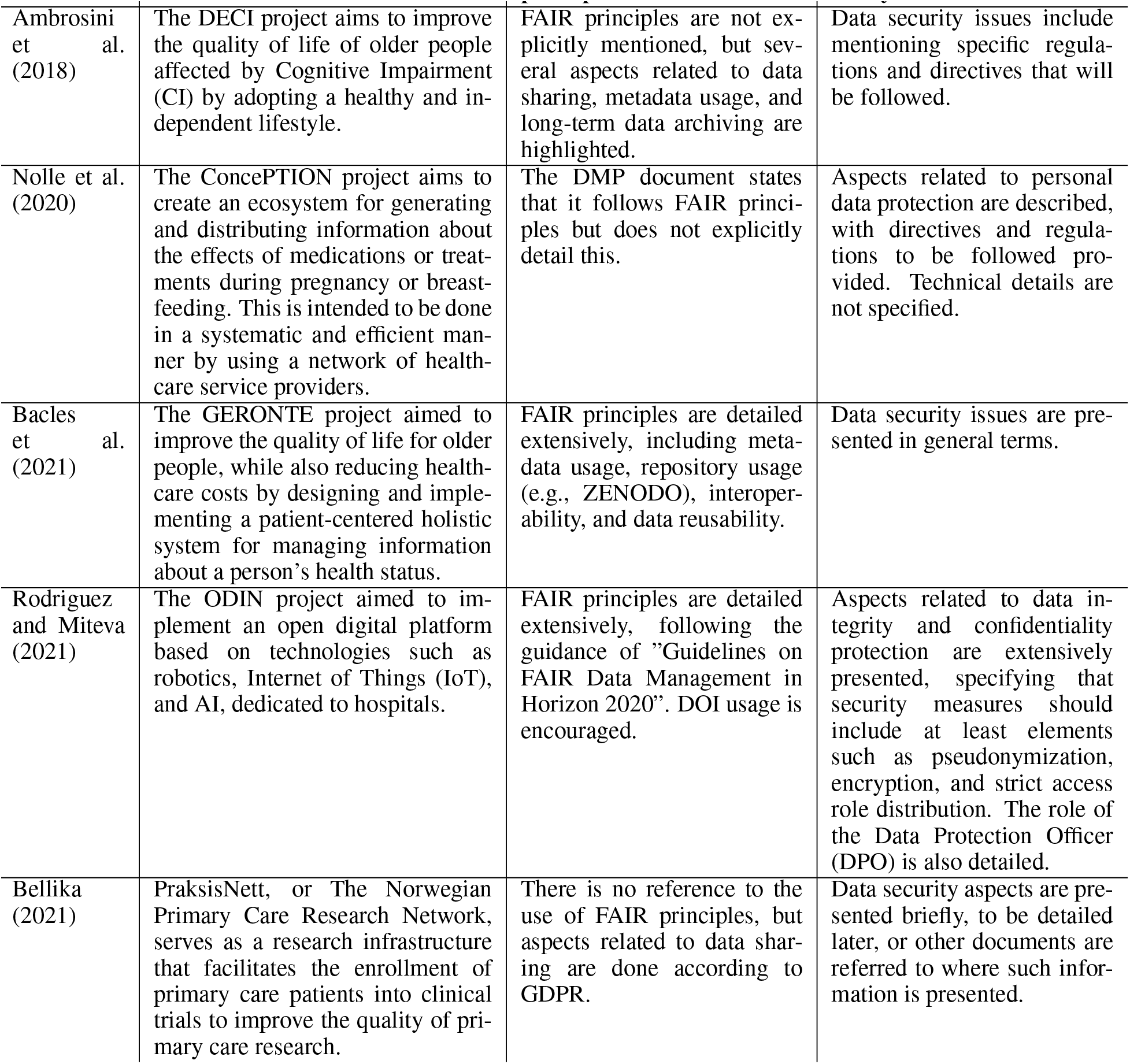
A brief survey on the usage of DMPs in healthcare-related research projects

| Reference | Purpose of the project | Aspects related to FAIR principles | Aspects related to cybersecurity |
| --- | --- | --- | --- |
| Valle et al. (2017) | The CrowdHEALTH project aims to integrate diverse information about a person's health status from multiple sources to support the definition and implementation of public health policies. | Due to the use of a large number of heterogeneous data sources, which need to be interoperable, and due to confidentiality constraints of most data sets, it was found that the FAIR model is not suitable for collecting information. However, a questionnaire was designed to collect information on the use of FAIR principles by a number of pilot sites participating in the project. | A question about data security was also introduced in the questionnaire for collecting information from pilot sites. |
| Ambrosini et al. (2018) | The DECI project aims to improve the quality of life of older people affected by Cognitive Impairment (CI) by adopting a healthy and independent lifestyle. | FAIR principles are not explicitly mentioned, but several aspects related to data sharing, metadata usage, and long-term data archiving are highlighted. | Data security issues include mentioning specific regulations and directives that will be followed. |
| Nolle et al. (2020) | The ConcePTION project aims to create an ecosystem for generating and distributing information about the effects of medications or treatments during pregnancy or breastfeeding. This is intended to be done in a systematic and efficient manner by using a network of health-care service providers. | The DMP document states that it follows FAIR principles but does not explicitly detail this. | Aspects related to personal data protection are described, with directives and regulations to be followed provided. Technical details are not specified. |
| Bacles et al. (2021) | The GERONTE project aimed to improve the quality of life for older people, while also reducing health-care costs by designing and implementing a patient-centered holistic system for managing information about a person's health status. | FAIR principles are detailed extensively, including meta-data usage, repository usage (e.g., ZENODO), interoperability, and data reusability. | Data security issues are presented in general terms. |
| Rodriguez and Miteva (2021) | The ODIN project aimed to implement an open digital platform based on technologies such as robotics, Internet of Things (IoT), and AI, dedicated to hospitals. | FAIR principles are detailed extensively, following the guidance of "Guidelines on FAIR Data Management in Horizon 2020". DOI usage is encouraged. | Aspects related to data integrity and confidentiality protection are extensively presented, specifying that security measures should include at least elements such as pseudonymization, encryption, and strict access role distribution. The role of the Data Protection Officer (DPO) is also detailed. |
| Bellika (2021) | PraksisNett, or The Norwegian Primary Care Research Network, serves as a research infrastructure that facilitates the enrollment of primary care patients into clinical trials to improve the quality of primary care research. | There is no reference to the use of FAIR principles, but aspects related to data sharing are done according to GDPR. | Data security aspects are presented briefly, to be detailed later, or other documents are referred to where such information is presented. |

### 2.1 FAIR Principles

The FAIR Guideline Principles aim to promote transparency, reproducibility, and re-usability of data. The acronym FAIR stands for Findable, Accessible, Interoperable, and Reusable, and it serves as a guide for data producers and managers.

In order to adhere to the FAIR principles, data should be findable, which means that it should have a unique and persistent identifier and richly described metadata that clearly identifies the data they refer to. Additionally, data should be deposited in a findable repository. Data should also be accessible, which means that it should be identified using standard and open protocols, and metadata should remain accessible (even if the data no longer exists).

Data should also be interoperable, which means that it should allow for exchange between platforms, be machinereadable, and refer to other sources of metadata when necessary. The clear identification of data sets is crucial for data integration. To be effective, these identifiers should be persistent, unique, and compliant with existing standards within the specific research community. This will ensure that the data set can be easily identified even if the physical repository or URL changes, which will facilitate data integration within specific infrastructures.

Lastly, data should be reusable, which means that it should be carefully and completely described, have a clear and accessible license, and comply with community-driven standards.

#### 2.1.1 Using Persistent Identifiers

FAIR principles underscores the importance of making data easily discoverable and machine-readable to enable automated attribution.

The first pillar of FAIR is Findable, which involves assigning a persistent identifier (PID) to the data for easy identification and indexing, allowing resources to be referred to using a single identifier that remains constant even if the resource’s URL changes. The PIDs should be registered and searchable by search engines (Wood-Charlson et al., 2022).

In addition, PIDs have various functions and benefits, as they allow for the linking of different research components, such as physical samples, instruments, organizations, digital objects, and individuals. PIDs also include metadata with standardized relationship terms that capture the connections between different research products and people.

These linkages can be used to automatically generate network graphs that connect researchers and their work to the wider publishing landscape. By doing so, researchers can discover connections they may not have been aware of previously. Most importantly, PIDs and their relationships enable researchers to receive credit for their contributions at a more detailed level than was previously possible.

One such digital identifier is the Digital Object Identifier (DOI), which not only uniquely identifies an object but also assigns a URL to the object’s metadata. This allows for increased data interoperability between humans and machines. By linking the DOI closely to the metadata, it becomes easier to share, discover, and integrate data sets across different platforms and communities.

### 2.2 Healthcare Data Security and Privacy

Data management is a critical part of the healthcare domain, involving the collection and management of large volumes of data from different sources.

Electronic health records (EHRs), medical imaging systems, and laboratory information systems are among the many means through which data is collected. Secure databases are used to store this data, which should to kept accurate, complete, and up-to-date, and advanced analytical tools and techniques like data mining, machine learning, and artificial intelligence are then used to analyze and visualize the data.

Healthcare data comes in different formats, with organized data being structured and easy to manage and analyze, while unorganized data like handwritten notes and free-text data can be difficult to interpret and use. EHRs provide healthcare professionals with access to a patient’s entire medical history, reducing the time in obtaining previous test results and improving care coordination between multiple healthcare providers. EHRs also help identify potential health risks, reduce medical errors, improve patient safety, empower patients to take an active role in their healthcare, and streamline administrative tasks, ultimately leading to cost savings.

Medical imaging is a vital part of modern healthcare, with various techniques like CT, MRI, X-ray, ultrasound, and others used depending on patient needs. The increasing use of medical imaging has led to the development of efficient systems like PACS, which allow for the storage and convenient access of medical images and reports, making it easier for healthcare professionals to access the information they need when treating patients.

However, data exchange relies on using structured data to retrieve medical images, which means that the data must be properly organized and tagged. The use of PACS systems has greatly improved the efficiency of medical imaging, making it easier for healthcare professionals to access and analyze the images they need to provide the best possible care for their patients.

In addition to EHRs and medical imaging, electronic medical records (EMRs) and other healthcare data components like personal health records (PHRs) have the potential to improve the quality, efficiency, and cost-effectiveness of healthcare while reducing medical errors. Data from various sources such as genomics-driven experiments, and the internet of things (IoT) can be analyzed to improve patient care, reduce healthcare costs, and improve healthcare outcomes.

#### 2.2.1 Healthcare Data Preparation and Sharing

The process of preparing data is crucial for accurate predictive models and reliable data mining techniques. Without proper preparation, processing raw data could require excessive computational resources, which is often not feasible in most cases. Data preparation is composed of two main steps, which are data filtering and data cleaning (El aboudi and Benhlima, 2018).

Data filtering aims to discard irrelevant data based on a specific criterion. On the other hand, data cleaning involves various procedures, such as noise reduction, normalization, and managing missing data. Medical records often contain noisy and missing data, and it is essential to eliminate noisy data and determine the values of missing data. Filling missing values inaccurately can lead to incorrect results and adversely affect the quality of the predictive model, therefore handling missing data must be carried out with utmost precision to prevent wrong decisions that may have serious consequences.

With large datasets, discarding irrelevant information that is not useful based on a defined criterion is necessary to optimize data processing and analysis. This can be achieved through various techniques such as feature selection, which selects only the most relevant features, or instance selection, which selects only the most informative instances.

Sharing data can have many benefits for scientific research and society as a whole. However, there are also several challenges associated with this process (Figueiredo, 2017). These challenges can be categorized into four levels: ethical/legal, cultural, financial, and technical. One of the primary ethical and legal challenges associated with sharing clinical trials and patient data is the need to de-identify the data to protect patient privacy.

However, even with de-identification, there is still a risk of re-identification for genomics and other related datasets. This can have significant implications for the ethical and legal use of this data. To address these challenges, solutions such as genome donation and open consent, combined with controlled data access and reliable data warehousing facilities, can help to protect patient privacy and ensure that data sharing is ethical and legal.

#### 2.2.2 Security Aspects Related to Healthcare Data

Smart healthcare systems use technology such as electronic health records (EHRs) and connected medical devices to improve patient care and outcomes. However, these systems also create new risks and vulnerabilities for sensitive medical information, including personal identifiable information (PII), health records, and medical images (Singh et al., 2021).

As a result, protecting the confidentiality, integrity, and availability of medical data is critical to maintaining patients’ trust and complying with legal and ethical obligations. Failure to secure medical data can lead to a range of negative consequences, including identity theft, medical fraud, and even harm to patients’ health. Therefore, it is essential to implement robust security measures and standards to safeguard medical data in smart healthcare systems. We review below several security techniques used for healthcare applications.

#### 2.2.3 Cryptography

Cryptography is a technique that involves converting information into an unreadable cipher, which can only be deciphered with a specific key. Cryptographic techniques provide essential security components such as confidentiality, integrity, and authentication.

For example, confidentiality refers to the protection of sensitive information from unauthorized access, while integrity ensures that the data is not tampered with or altered. Authentication verifies the identity of the user accessing the data.

Cryptography can be used to encode the data into an unreadable alpha-numeric text. This means that even if an unauthorized person gains access to the data, they would not be able to read or interpret it without the decryption key. This is an important feature of cryptography in ensuring the privacy and security of sensitive digital data, such as medical records.

Two main cryptographic techniques used in practice are asymmetric and symmetric key-based encryption schemes, which are briefly described below.

Asymmetric encryption uses a public-private key pair to encrypt and decrypt messages. Data encrypted with the public key can only be decrypted with the corresponding private key. This technique provides a high level of security, but the maintenance, distribution, and exchange of keys can be challenging.

Symmetric encryption, on the other hand, uses only one key for both encryption and decryption. While this technique simplifies key management, it faces the challenge of man-in-the-middle attacks, where an attacker intercepts and modifies the communication between two parties.

In addition to encryption, cryptographic techniques can also use hashing and digital signatures to verify ownership authentication and detect tampering or modifications to medical images. Hashing involves generating a unique identifier for each image, which can be used to verify ownership and detect any changes to the image. Digital signatures, on the other hand, involve using a mathematical algorithm to create a unique digital signature that can only be generated by the owner of the image.

For instance, Al-Haj et al. (2015) present two algorithms that use cryptography to ensure the confidentiality, authenticity, and integrity of DICOM images. Unlike other cryptographic schemes and the DICOM standard, the proposed algorithms provide these security features for both the header data and the pixel data of the images. The algorithms use strong cryptographic functions and symmetric keys, as well as hash codes. These features are implemented in the algorithms to ensure secure storage and transmission of DICOM images

Another example is a highly robust method for hiding information in images, which was presented by Arunkumar et al. (2019). This technique uses a combination of Redundant Integer Wavelet Transform (RIWT), Discrete Wavelet Transforms (DCT), and Singular Value Decomposition (SVD), as well as the logistic chaotic map. By using RIWT, the proposed method achieves reversibility, shift invariance, and robustness. SVD and DCT are used to achieve a high level of imperceptibility by embedding data in singular values. The logistic chaotic map is used to encrypt secret medical images and enhance the robustness of the technique.

#### 2.2.4 Biometrics

Biometrics is a scientific method that uses physical, chemical, or behavioral characteristics to establish an individual’s identity with a high degree of accuracy and reliability (Jain et al., 2008). Biometric technology has become an increasingly popular way to verify or identify individuals in various applications such as security, access control, identification, and authentication.

Through the use of biometric systems that capture and analyze biometric data, unique traits such as fingerprints, iris texture, facial features, and voice patterns can be used to verify the identity of an individual. This method is particularly advantageous as biometric traits are difficult to replicate or forge, making it a secure and reliable means of identification.

A biometric system typically operates in one of two modes: verification mode or identification mode. Verification mode is used to validate the identity of an individual by comparing their captured biometric information with their own stored biometric template in the database. On the other hand, identification mode is used to identify an individual by searching the biometric templates of all users in the database for a match.

For example, Ramli et al. (2016) propose a biometric system that is based on Electrocardiographic (ECG) signals, which reflect the mechanical movement of the heart. This modality is chosen because it cannot be faked, unlike fingerprints, which can be fooled with fake fingers, and faces, which can be extracted using photos. Voice can also be conveniently imitated. ECG signals contain unique physiological information that makes them a promising authentication technology. A portable ECG detection kit is developed for data acquisition, and a wearable bracelet is used for personal system login. Wavelet transform algorithm is used for feature extraction, while Support Vector Machine (SVM) is employed for the classification process. The prototype has been successfully tested and shows promising results for authentication purposes.

However, several issues have been identified with biometric-based authentication approaches. These issues include the need to consider accuracy rate, security, cost, robustness against attacks, computational time, and scalability of the system during development, which can be challenging for researchers to maintain all of these factors simultaneously. The security and privacy of biometric templates are also major concerns. Additionally, performance, acceptability, and circumvention are other important issues to consider for practical biometric systems. Maintaining a large database of biometric data is also necessary, which can be a significant challenge. Finally, the computational complexity of these systems is high, which can pose additional difficulties for implementation.

#### 2.2.5 Digital Watermarking

Several authors have developed digital watermarking techniques to provide copyright protection and content authentication for health data (Soni and Kumar, 2020; Kaur et al., 2010; Giakoumaki et al., 2006; Rizzi et al., 2020). This technique involves inserting various types of medical and patient information into medical images to establish ownership and maintain integrity while preserving the visual quality of the cover data.

Medical image watermarking also offers protection against tampering, access control, non-repudiation, indexing, and reduces memory and bandwidth requirements. Research has shown that imperceptibility, robustness, and capacity are fundamental requirements of any watermarking technique.

For instance, iris identification is considered the most reliable form of biometric identification, but the iris images collected for identification purposes can be stored in databases that may be vulnerable to hacking by intruders. To prevent these databases from being tampered with by adding watermark text, a hybrid method has been proposed by Mothi and Karthikeyan (2019) that combines Wavelet Packet Transform (WPT) with cryptography. The proposed approach uses WPT to segment the iris image and locate the minimum energy band where the watermark text containing the owner’s personal information is embedded. Once the watermarking is completed, the watermarked image is encrypted using a cryptographic key, effectively preventing both the image and the watermark text from being tampered with in an efficient manner.

A robust and secure mechanism is needed to transfer medical images over the internet, and Sharma et al. (2015) have proposed a method for watermarking which is based on two popular transform domain techniques, discrete wavelet transforms (DWT) and discrete cosine transform (DCT). During the embedding process, the cover medical image is divided into two parts, region of interest (ROI) and non-region of interest (NROI).

Multiple watermarks in the form of image and text are embedded into ROI and NROI parts of the same cover media object, respectively, for identity authentication purposes. To enhance the security of the text watermark, the Rivest-Shamir-Adleman (RSA) encryption technique is applied before embedding, and the encrypted EPR data is then embedded into the NROI portion of the cover medical image.

In today’s healthcare systems, medical equipment produces digital images that need to be securely stored and exchanged to protect patient privacy and image integrity. Reversible watermarking techniques can be used for this purpose. To this end, Abd-Eldayem (2013) proposes a security technique based on digital watermarking and encryption for Digital Imaging and Communications in Medicine (DICOM) to provide patient authentication, information confidentiality, and integrity using reversible watermarking.

To ensure integrity at the sender side, a hash value based on encrypted MD5 is generated from the image. To maintain reversibility, an R-S-Vector is obtained from the image and compressed using Huffman compression. To provide confidentiality and authentication services, the compressed R-S-Vector, hash value, and patient ID are concatenated to form a watermark, which is then encrypted using the AES encryption technique and embedded in the medical image.

The proposed technique achieves high imperceptibility, invisibility, and transparency with excellent efficiency. The watermark extracted at the receiver side is identical to the embedded watermark at the sender side, ensuring total reversibility without affecting the quality of the original image.

## 3 Materials and Methods

For this systematic review, we employed the PRISMA methodology (Page et al., 2021). We searched the PubMed and OpenAlex (Priem et al., 2022) databases for relevant articles using the query service provided by Lens.org (Jefferson et al., 2021).

The methodology is illustrated in Figure 1. Relevant articles were identified through the query “healthcare AND data AND fair,” which was applied to the abstracts of the records.

**Figure 1:**
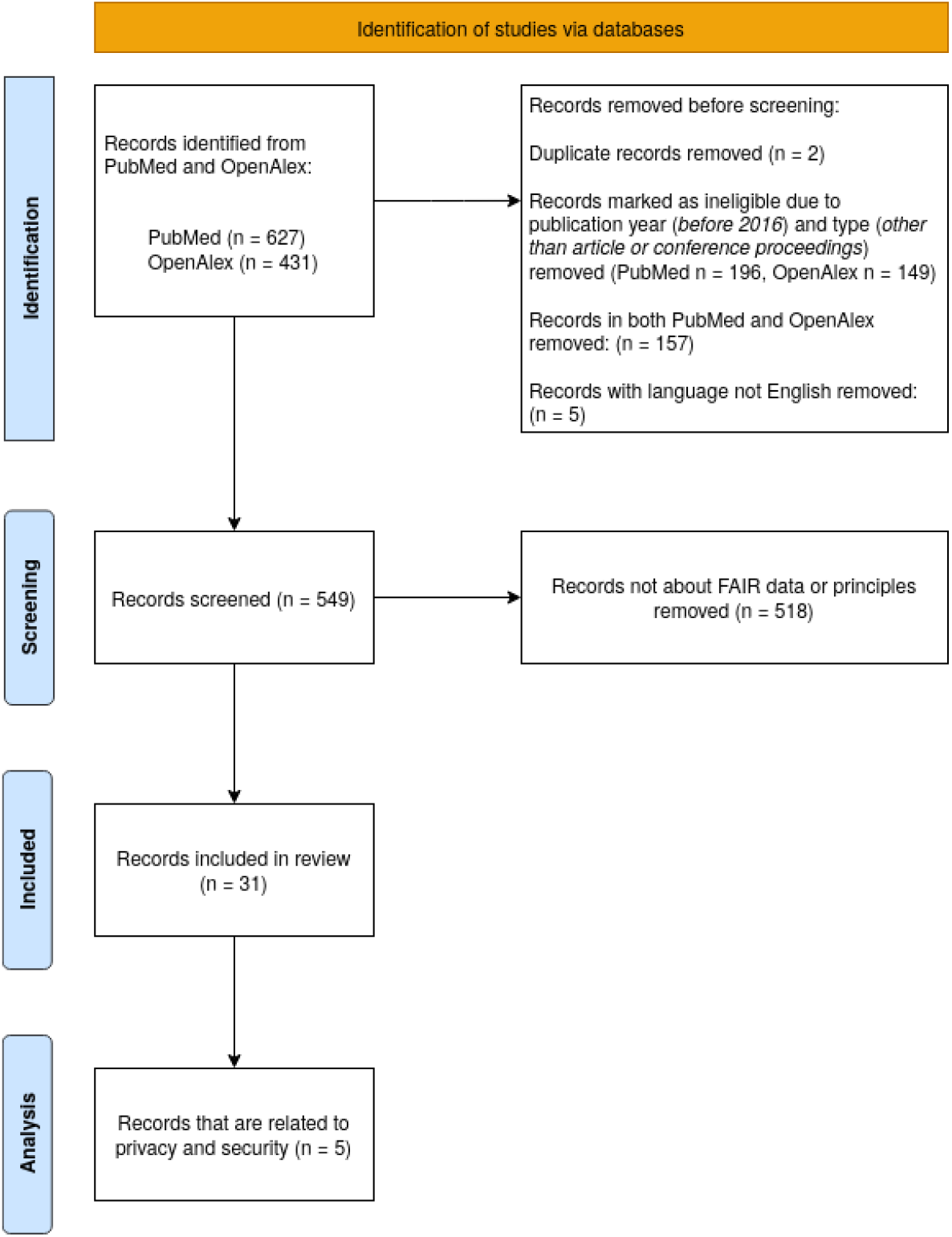
Flowchart of the collection and processing of records from PubMed and OpenAlex databases

We identified a total of n = 627 records in PubMed and n = 431 records in OpenAlex.

Eligibility criteria for including articles in the review were established as follows. The publication date of the articles must be later than 2016 (inclusive), as the first references mentioning the FAIR principles were published that year. We selected only articles that underwent a peer-review process, i.e., articles published in a journal or conference proceedings.

Articles not written in English (n = 5) were excluded. Duplicate records were also removed, and those appearing in both databases were included only once. Thus, in the filtering stage, n = 549 records were selected. Articles were automatically filtered by searching for the string “FAIR” within the abstracts, and then manual identification was performed for the records that did not refer to the FAIR principles. Ultimately, a total of n = 31 articles were included in the review.

Subsequently, a categorization of these articles was considered, given that there is no universally recognized taxonomy for research articles that focus primarily on FAIR principles and their application in the healthcare domain. However, these articles can be grouped based on the research subject they concentrate on. In this regard, the following three categories were considered:

- Overview and adoption of FAIR principles: This category refers to topics such as general reviews and surveys, case studies, as well as challenges and barriers to adoption. The objective of this category is to provide a comprehensive understanding of the FAIR principles, their acceptance within the research community, and the factors influencing their implementation.
- FAIRification of data: This category encompasses aspects such as data management plans and best practices, methodologies for data curation and quality control, and long-term data preservation and archiving, including security and privacy considerations. The focus of this category is to highlight the processes and techniques involved in ensuring data adheres to the FAIR principles, promoting its reusability, accessibility, and interoperability.
- Infrastructure and tools: This category includes subjects such as data repositories and platforms, metadata standards and ontologies, as well as software tools and services for FAIR data management. The emphasis here is on the technological solutions and frameworks that support the FAIRification of data, enabling researchers to effectively manage and share their data in adherence to the FAIR principles.

For the classification of the articles selected for the review, we used the InstructGPT model (Ouyang et al., 2022) provided by OpenAI, with the following prompt:

~~~
“You need to decide in which category the following text falls into according to its content.
    Possible choices are:
‘Overview and Adoption of FAIR Principles’ (that could include general reviews and surveys, case    studies, guidelines, challenges and barriers to adoption),
‘FAIRification of data’ (that could include methodologies for data storage, access, curation,
    preservation, archiving, security, privacy),
‘Infrastructure and Tools’ (that could include data repositories and platforms, metadata standards
    and ontologies).\n\nText: {Abstract}
\nCategory: “
~~~

The InstructGPT model used was *text-davinci-003*, and the Jupyter notebook and all data used for analysis in this study are available in a Binder repository (Stanciu, 2023).

For two of the categories, the review was performed manually, whereas for the third category, the analysis was done with the InstructGPT language model using the following prompt:

~~~
“Explain the role of FAIR principles in the following text. Use only three sentences. Do not tell
what the FAIR principles are.\n\nText: {Abstract}
\nSummary: “
~~~

The combination of human review and the InstructGPT tool aimed to balance the advantages of both methods. While human reviewers brought their expertise and ability to understand the context, the InstructGPT tool offered a streamlined, objective, and consistent analysis. By employing these two distinct approaches, the study aimed to provide a comprehensive and accurate evaluation of the articles across all three categories.

Finally, a further refinement of the initially selected 31 articles for review was conducted with the aim of identifying which of these specifically address the topics of security or data privacy. As a result, a total of n = 5 articles were identified as relevant to these themes.

For the analysis and visualization of the results, the VOSviewer tool (Perianes-Rodriguez et al., 2016) was employed.

## 4 Results

The articles selected for review were grouped into three categories according to the topics addressed. Their distribution was done automatically using the InstructGPT model, and the results of this preliminary automated categorization are presented in Table 2.

**Table 2:** Distribution of articles by categories according to the subjects addressed

| Topic | # of articles |
| --- | --- |
| Overview and adoption of FAIR principles | 18 |
| Infrastructure and tools | 8 |
| FAIRification of data | 5 |

Out of the 31 studies selected for analysis, it was found that one study did not have FAIR principles as its topic, namely Scherer et al. (2022) discuss the implementation of a support system for recruiting participants in clinical trials based on fast healthcare interoperability resources (FHIR) specifications.

A synthesis of the selected articles for the fields of ‘FAIRification of data’ and ‘Infrastructure and tools’ is presented below.

Ogundepo et al. (2020) presents a preliminary assessment of a dataset regarding the COVID-19 pandemic in Nigeria, which was generated by integrating heterogeneous sources and using FAIR principles.

Queralt-Rosinach et al. (2022) discusses the experience of applying FAIR principles in managing hospital data during the COVID-19 pandemic, highlighting the main difficulties and opportunities that arose. The FAIRification process of the data was based on the use of ontologies for data and metadata.

The study of Urwin et al. (2022) presents the experience gained within the CO-CONNET project dedicated to the COVID pandemic response in the UK, as one project objective was to transform a dataset according to FAIR principles.

Folorunso et al. (2022) presents a data flow model for machine learning algorithms, which was implemented according to FAIR principles. Their study also discusses the aspects related to the FAIRification process of the data using the dataset presented by Ogundepo et al. (2020).

A new data representation model, ‘term sets,’ is presented in the study Williams et al. (2019), providing transparency and reproducibility of research results achieved according to FAIR principles.

The study of Sinaci et al. (2023) describes a methodology for transforming data according to FAIR principles, and the study of Estupiñán (2022) presents the federated PHIRI infrastructure, which includes four use cases in which data hubs represent the nodes of the federated network.

Blomberg and Lauer (2020) presents the experience accumulated within the Elixir research infrastructure, where FAIR principles were applied.

The study of Deist et al. (2020) presents the Personal Health Train (PHT) infrastructure, and the implementation is according to privacy-by-design principles. The connected FAIR data sources are used to train distributed machine learning algorithms.

Sollis et al. (2023) presents the NHGRI-EBI GWAS catalog, which was implemented as a knowledge base and is available as FAIR resource.

The study of van der Velde et al. (2022) describes a metadata scheme for FAIRifying genomic data. This was achieved using ontologies such as NCIT, DUO, and EDAM. This scheme is represented in YAML format and can be transferred within a template using JSON or RDF formats to support genomic data sharing.

In the study of Welten et al. (2022), a system for detecting breast cancer using a distributed infrastructure for data analysis is presented. In this regard, a detailed analysis of the possibilities of applying FAIR principles is carried out.

Among these studies, Welten et al. (2022); Deist et al. (2020); Sinaci et al. (2023) also address issues related to data security and privacy.

In addition, Daniel and Choquet (2017) presents an overview of the principal research subjects from 2016 in clinical research informatics (CRI), addressing aspects related to data privacy, and the study of Kush et al. (2020) discusses aspects related to data security and privacy within the FAIR data-sharing process, highlighting the standardization effort using common data elements (CDEs).

Subsequently, for the articles grouped in the category ‘Overview and adoption of FAIR principles’, a review was conducted using the InstructGPT tool. The response generated by the *text-davinci-003* model was analyzed, and its accuracy level was assessed.

### Summarization using InstructGPT

Kalendralis et al. (2021): The FAIR principles are discussed in this article in relation to the use of artificial intelligence in radiotherapy. The article outlines the need for and usage of the FAIR principles, and provides guidance and potential applications of FAIR to various radiotherapy stakeholders. It emphasizes the importance of medical physicists in this process.

Assessment: The response is accurate.

Wang and Williams (2022): The FAIR principles are used as a framework to discuss the structural reasons why artificial intelligence has not yet been able to make full use of existing datasets. The principles are used to identify barriers preventing artificial intelligence from making use of existing datasets, and to suggest ways to enable progress. The Brain Tumour Data Accelerator is an example of how the FAIR principles can be used to integrate fragmented patient data into an enriched dataset.

Assessment: The response is accurate.

Harrow et al. (2022): The FAIR principles are being used to enable a shift from an application-centric to a data-centric perspective in the life sciences. This shift is necessary to maximize the value of data generated from research, clinical trials, and real-world healthcare data. Use cases for FAIR implementation are being considered alongside assessment of data quality to achieve this goal.

Assessment: The response is accurate.

McCarthy (2016): The FAIR Health non-profit corporation conducted an analysis of privately billed healthcare claims to show the rise of opioid dependence in the United States. The FAIR principles ensure that the data collected is accurate, transparent, and accessible to all stakeholders. This data is then used to inform government agencies, insurers, providers, and consumers of the current opioid epidemic in the US.

Assessment: The response is accurate.

Kush et al. (2020): The FAIR principles are a set of guidelines that aim to ensure that data is Findable, Accessible, Interoperable, and Reusable. The text discusses the challenges of data sharing and interoperability, and how the lack of standards and traceable semantics can make it difficult to adhere to the FAIR principles. The text also suggests that common data elements (CDEs) could be used to standardize data collection and enable responsible data sharing that would help to adhere to the FAIR principles.

Assessment: The response is accurate.

Biloborodova et al. (2021): The FAIR principles are used in this paper to ensure data reuse and improve image classification performance in imbalanced datasets. The proposed framework is tested on an acne disease dataset and is shown to outperform the baselines, with up to 5% improvement in image classification. The FAIR principles are essential for ensuring data reuse and accuracy in medical image classification and diagnosis.

Assessment: The response is accurate.

Belien et al. (2022): The FAIR principles are used as the basis for this study to bridge gaps in the management and sharing of NGS data. The analysis is part of a programme to develop an instruction manual for FAIR genomic data in clinical care and research. The gap analysis is a possible contribution from the Netherlands to the European 1+ Million Genomes Initiative.

Assessment: The response is accurate.

Daniel and Choquet (2017): The FAIR Guiding Principles are used to evaluate the risk of privacy breaches in releasing genomics datasets. They are also used to create phenotype models via a machine learning process and to securely connect ResearchKit compatible applications to the widely-used clinical research infrastructure Informatics for Integrating Biology and the Bedside (i2b2). The FAIR principles are essential for ensuring the safe and secure use of data in the field of Clinical Research Informatics.

Assessment: The response is not relevant and is not accurate.

Kodra et al. (2018): The FAIR principles are used as a framework to improve the quality of RD registries. This includes aspects of governance, data and information, infrastructure, documentation, training, and quality audit. The list of recommendations is intended to be used by established and new RD registries, and a toolkit is being developed to enable continuous assessment and improvement.

Assessment: The response is accurate.

Hulsen et al. (2023): The FAIR principles provide guidelines to make data findable, accessible, interoperable and reusable to the research community. Federated learning, standards and ontologies are useful to improve robustness of artificial intelligence algorithms working on big data. Big data and artificial intelligence also offer opportunities for laboratories to optimize the productivity of the laboratory, the quality of laboratory results and ultimately patient outcomes.

Assessment: The response is accurate.

Hutton et al. (2023): The FAIR principles are used in this analysis to obtain in-network costs from the FAIR Health Consumer online database. These costs are then used to calculate the overall cost burden per patient for all surgeries, diagnostics, and treatments. The results of this analysis show that Chronic Pelvic Pain has a large cost burden of $29,951 for in-network services. Assessment: The response is accurate.

Bhatia et al. (2020): The FAIR principles are used to improve the findability, accessibility, interoperability, and reusability of scholarly data. In this case study, the FAIR principles were used to develop a search tool, EDCat, to improve the FAIRness of HCUP-SEDD. The publication of data dictionaries should be encouraged to further improve the FAIRness of datasets.

Assessment: The response is accurate.

Adler-Milstein et al. (2019): The FAIR principles are essential for healthcare delivery organizations to be able to spread data science discoveries at scale. The principles ensure that knowledge is findable, accessible, interoperable, and reusable. The conference discussed the need for healthcare delivery organizations to develop new organizational capabilities across three domains in order to engage with computable knowledge management. These capabilities must be based on the FAIR principles in order to be successful.

Assessment: The response is mostly correct, but it does not provide the relevant information.

Klofenstein et al. (2021): The FAIR principles are used to ensure that data is Findable, Accessible, Interoperable, and Reusable. This text discusses how the mapping of a generic metadata schema to the HL7® Fast Healthcare Interoperability Resources® (FHIR)® standard can improve data FAIRness and widen analysis possibilities. The mapping results showed that 94% of the items could be mapped to FHIR, with 61%, 57%, and 52% of the items available as standard resources for studies, questionnaires, and documents, respectively. This demonstrates the potential of FHIR to reduce data ambiguity and foster interoperability.

Assessment: The response is accurate.

Jefferson et al. (2022): The FAIR principles are applied in the text to ensure that UK COVID-19 data is Findable, Accessible, Interoperable and Reusable. This is achieved through a federated platform which enables researchers to discover data and meta-analysis, as well as detailed analysis with data governance approvals. Finally, the FAIR principles allow for rapid, robust research to be conducted in a de-identifiable and safe way.

Assessment: The response is accurate.

Puttmann et al. (2022): The FAIR principles are essential for health data to be internationally Findable, Accessible, Interoperable and Reusable. The Dutch National Intensive Care Evaluation (NICE) quality registry adopted the Observational Medical Outcomes Partnership Common Database Model (OMOP CDM) to achieve a FAIR database. Through communication, research and trial-and-error, solutions were found to help other healthcare institutions FAIRify their databases.

Assessment: The response is accurate.

Bourgeois (2022): The FAIR principles are used to ensure that data generated from pediatric clinical trials is Findable, Accessible, Interoperable, and Reusable. This allows external investigators to build on prior work and encourages data sharing and reuse. The authors suggest that a data-driven approach should be used to monitor activities and direct modifications to research infrastructures, policies, and investment strategies.

Assessment: The response is mostly accurate.

Oladipo et al. (2022): The FAIR principles are used to make digital data Findable, Accessible, Interoperable, and Reusable. A digital initiative has been developed to foster professional skills in data stewardship through effective knowledge communication. The FAIR Data Management course offers 6 short on-demand certificate modules to certify students as FAIR data scientists and qualified to serve as both FAIR data stewards and analysts.

Assessment: The response is accurate.

### 4.1 Vizualization of Term Co-Occurrences

VOSviewer was employed to create detailed graphs that illustrate the co-occurrence of terms found within the abstracts of the selected articles. By analyzing these graphical representations, the relationships and connections between various concepts and ideas can be better understood. A specific example of this type of visualization, showcasing the complex interplay of terms, can be seen in Figure 2. In a similar manner to the previous example, Figure 3 showcases a visualization that represents the co-occurrence of keywords found within the selected articles. The purpose of this visualization is to provide a clear and concise overview of how frequently certain keywords appear together, revealing thus patterns and relationships between them.

**Figure 2:**
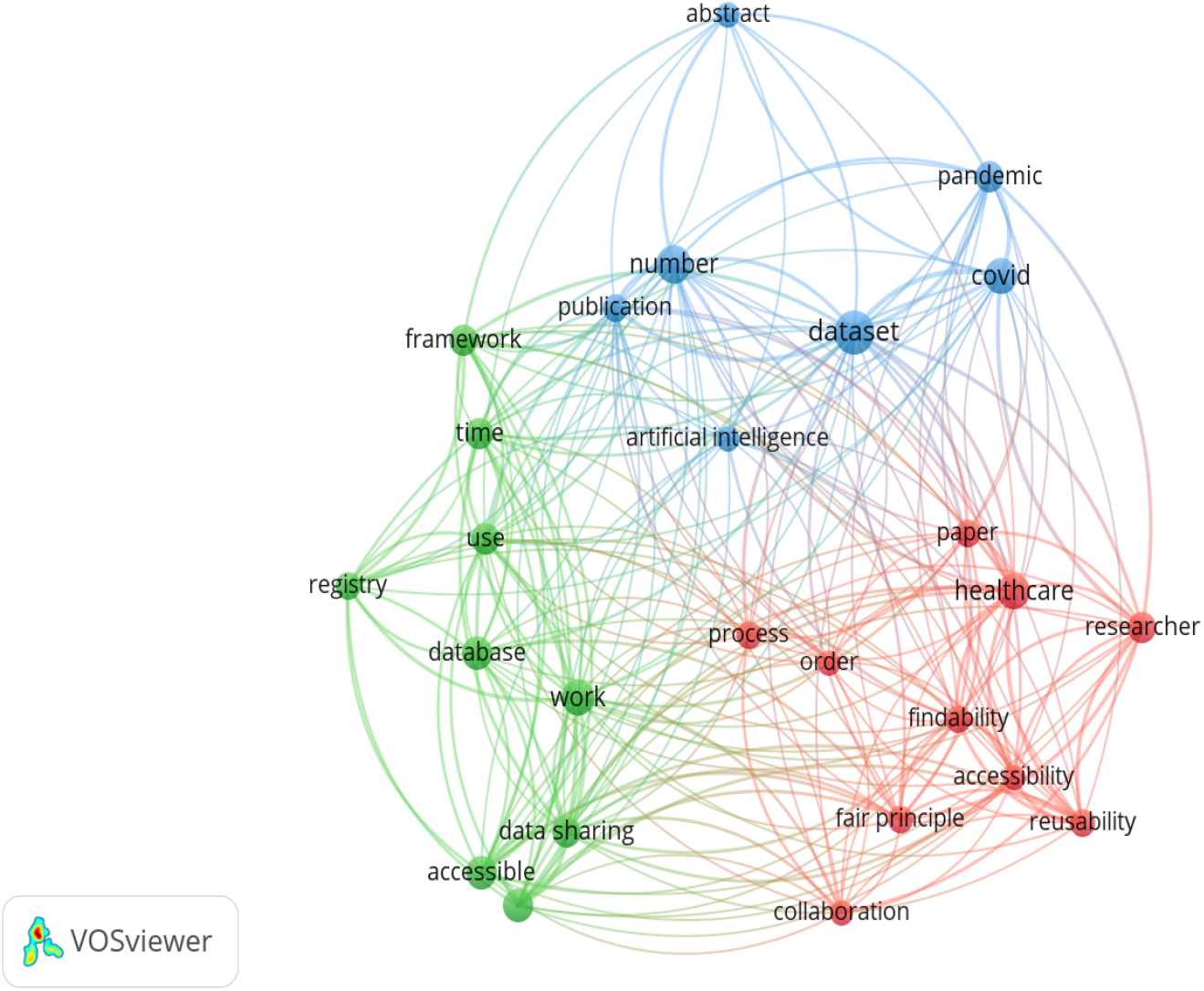
Co-occurrence of terms present in Abstract

**Figure 3:**
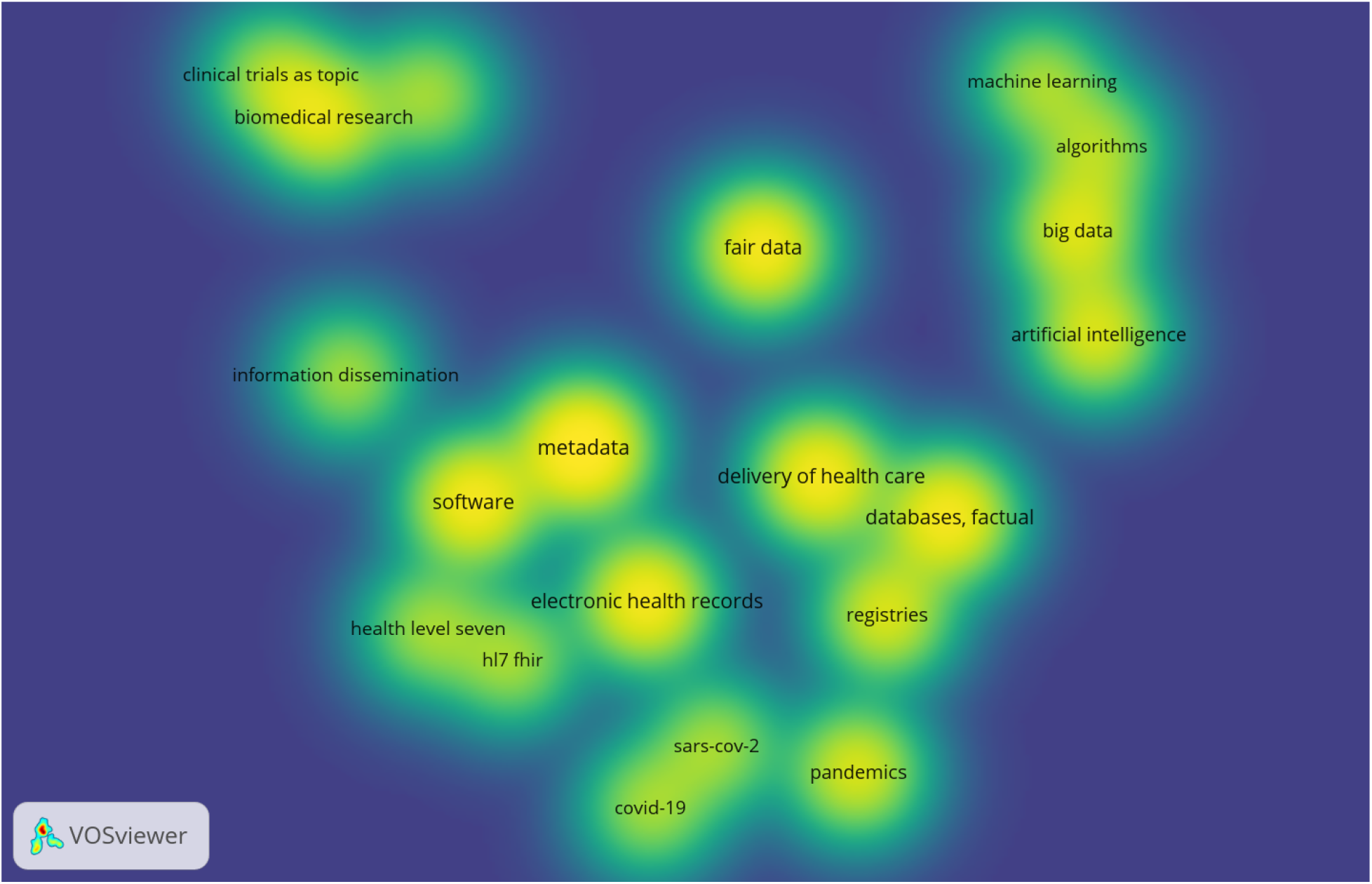
Keywords co-occurrence map

To achieve this, the figure employs a density visualization technique, which effectively conveys the distribution and intensity of keyword co-occurrence by using variations in color or shading. As a result, this method allows for a more intuitive understanding of the data, making it easier to discern the formation of multiple clusters.

These clusters, in turn, can be indicative of different themes or topics emerging from the analyzed articles. By identifying these groupings, one can gain valuable insights into the relationships between various concepts and ideas, which may help to highlight areas of interest for further exploration.

Thus, a cluster can be observed consisting of elements such as artificial intelligence, big data, algorithms, and machine learning. Another cluster can be considered to be the one formed by the terms metadata and software. These themes can be considered subjects of significant current interest and have an excellent perspective for further development, making them important to consider when using FAIR data.

### 4.2 Discussion

The InstructGPT tool, developed by OpenAI, has shown promising results for classifying and reviewing the articles chosen for the survey. However, it is essential to exercise caution when applying this technology, as some limitations have been observed. While the tool has provided accurate answers in most instances, there have been cases where the information presented was either irrelevant or partially incorrect.

This highlights that, despite the impressive performance of AI tools like InstructGPT, they are not perfect. These models rely on vast amounts of data and advanced algorithms to generate responses. However, they may still produce errors or lack the nuanced understanding of a specific domain that an expert possesses. Consequently, it is still necessary to have a domain expert to ensure the accuracy and relevance of the generated answers.

Furthermore, the expert’s role in the process includes verifying the correctness of the information provided and identifying potential biases or misconceptions that may arise from the model’s training data. By combining the expertise of human professionals with the computational power of tools like InstructGPT, a more reliable and efficient approach to the specific task can be achieved.

## 5 Conclusion

A data management plan is required for a research project, as it involves producing and analyzing data. It includes a detailed description of the procedures on how data will be generated, collected, processed, stored, shared, and distributed throughout the research project’s life cycle. In order to create an effective DMP, it is essential to align with the FAIR principles for research data management.

In this study, we have provided an analysis of DMPs, and have discussed the case of healthcare data management. Special consideration was given to the security-related aspects, which are paramount in the health sector. Further, a systematic survey was undertaken to identify current trends in the field and the solutions that can be considered for inclusion within a healthcare-focused DMP.

Also, the use of tools like InstructGPT offers significant advantages in terms of efficiency and scalability for various tasks, including article classification and review. However, it is essential to acknowledge their limitations and involve domain experts to ensure accurate and relevant results.

## Data Availability

All data produced are available online at  https://zenodo.org/record/7857251

## Notes

### Competing Interest Statement

The authors have declared no competing interest.

### Funding Statement

This study did not receive any funding.

### Summary of Updates

Section 3 was expanded to include the use of InstructGPT to review a group of articles, and Section 4 was updated accordingly with the analysis and the assessment. In Section 4, two sub-sections were introduced - A discussion and a part dedicated to the Visualization of the Term Co-Occurences.

